# Remdesivir Efficacy in Coronavirus Disease 2019 (COVID-19): A Systematic Review

**DOI:** 10.1101/2020.06.15.20131227

**Authors:** Amirhossein Roshanshad, Alireza Kamalipour, Mohammad Ali Ashraf, Romina Roshanshad, Mohammad Reza Akbari

## Abstract

**Background:** Researchers are working hard to find an effective treatment for the new coronavirus 2019. We performed a comprehensive systematic review to investigate the latest clinical evidence on the treatment efficacy and safety of Remdesivir in hospitalized patients with COVID-19.

**Methods:** We performed a systematic search of the Pubmed, Embase, Web of Science, Google scholar, and MedRxiv for relevant observational and interventional studies. Measured outcomes were mortality rates, improvement rates, time to clinical improvement, all adverse event rates and severe adverse event rates.

**Results:** 3 RCTs and 2 cohorts were included in our study. In 2 cohort studies, patients received Remdesivir for 10 days. 2 RCTs evaluated 10-day treatment of Remdesivir efficacy versus placebo group and the other RCT compared its 5-day regimen versus 10-day regimen. Visual inspection of the forest plots revealed that Remdesivir efficacy was not much different in reducing 28-day mortality versus 14-day mortality rates. Besides, 10-day treatment regimen overpowers 5-day treatment and placebo in decreasing time to clinical improvement. All adverse event rates did not have significant difference; however, severe adverse event rate was lower in 5-day Remdesivir group compared to 10-day and placebo groups.

**Conclusion:** 5-day course of Remdesivir therapy in COVID-19 patients is probably efficacious and safe and patients without invasive mechanical ventilation benefit the most. Treatment can be extended to 10 days if satisfactory improvement is not seen by day 5. Most benefits from Remdesivir therapy take place in the first 14 days of the start of the treatment.

## Background

Since the beginning of the coronavirus disease of 2019 (COVID-19) in Wuhan, it has infected more than 7.1 million people and caused more than 400000 deaths worldwide(1). The rapid spread of the virus has stricken global health and the economy(2). Therefore, researchers are working hard to find an effective treatment for the new coronavirus 2019

Currently, there is no established treatment, and supportive care is the only management in these patients(3). Many drugs tend to show up in many in vitro drug screens. However, their clinical effect is still under investigation. The medical community is actively trialing repurposed and novel medications to find an effective treatment(4). Drugs such as Remdesivir have been the subjects of recent studies in this situation.

Remdesivir (also known as GS-5734) is an adenosine analog, firstly developed against the Ebola virus in 2017(5). Recent studies showed promising results against the RNA coronaviruses (SARS-CoV and MERS-CoV)(6, 7). A study reported its clinical benefits in rhesus macaques infected with SARS-CoV-2(8).In addition, other studies revealed its in vitro efficacy against SARS-CoV-2 in human cell lines(9). However, Remdesivir effect does not necessarily translate into in vivo efficacy. Some studies investigated Remdesivir clinical efficacy in human patients; however, most of them were small groups, underpowered and inconclusive.

On May 1, 2020, FDA issues emergency use authorization for the administration of Remdesivir in COVID-19 patients(10). However, its efficacy is still controversial among experts. Therefore, it is essential to gather all available clinical studies to provide unified evidence on its efficacy.

Here, we performed a comprehensive systematic review to investigate the latest clinical evidence on treatment efficacy and safety of Remdesivir in hospitalized patients with COVID-19.

## Methods

### Protocol

In this study, we adhered to the recommendations provided by the Preferred Reporting Items for Systematic Reviews and Meta-Analyses (PRISMA) statement 2015(11).

### Literature search

We performed a systematic search of the MEDLINE(PubMed), EMBASE, and Web of Science databases with the MeSH and non-MeSH terms and the keywords of ‘coronavirus’ OR ‘COVID-19’ OR ‘SARS-COV-2’ OR ‘2019-ncov’ AND ‘GS-5734’ OR ‘Remdesivir’ on May 29, 2020. Google scholar and MedRxiv were also searched with these keywords manually to retrieve gray literature. Publication year were limited to 2020. Details about search strategy and the keywords are presented in Additional file. Except for the time, no other restriction was considered for our search. We also looked into the references of the included papers for more relevant studies.

Screening was done independently by M.A and A.R. Firstly, duplicated retrieved search results were identified and excluded. We screened the titles and the abstracts of the papers to exclude irrelevant studies. Accordingly, search results were categorized into three categories including included, excluded, and unclear. Then, Full texts of the retrieved studies were reviewed for final inclusion. Any disagreement was discussed in our project team.

### Selection criteria

#### Inclusion criteria

Original articles comparing treatment efficacy and safety of COVID-19 patients with Remdesivir and placebo were included. Also, studies evaluating treatment outcomes of COVID-19 patients with Remdesivir in groups with different duration of treatment or with dissimilar disease severity were included.

#### Exclusion criteria

Review articles, duplicate studies, meeting abstracts, letters and editorials, case reports and case series, and studies with no original information about Remdesivir therapy were excluded. In vitro and animal studies were also excluded.

### Data extraction

Author’s name, study design, number of patients, inclusion criteria, duration of illness before Remdesivir therapy, arms or subgroups of the patients, improvement and/or mortality rates, viral load changes and adverse events were extracted.

Two authors, A.R and R.R, performed literature search, screening and inclusion of the studies, and data extraction independently, and disagreements were discussed with another author, MRA, and resolved.

### Risk of bias assessment

We use Cochrane Risk of Bias Tool (12) and Newcastle-Ottawa Scale(13) for assessing risk of bias of randomized controlled studies and observational studies, respectively. Two authors, A.R and R.R independently assessed the risk of bias with the mentioned tools and M.A rechecked it.

### Statistical analysis

Results of the final studies were reported separately in a narrative way. Measured outcomes were mortality rates, time to clinical improvement, any adverse events, severe adverse events, and improvement rates. We used Review Manager 5.3 to draw forest plots of these outcomes. I^2^ was considered as the indicator for heterogeneity. Random effect model was used when I^2^ exceeds 50%. Mantel–Haenszel odds ratio (OR) and mean difference (MD) were used for reporting effects of dichotomous and continuous outcomes, respectively. Due to the great heterogeneity seen in these forest plots, meta-analysis was not possible. However, forest plots comparing measured outcomes in different subgroups were drawn. The day of measuring the outcomes (28-day vs. 14-day), types of the studies (RCT vs. Cohort), and design of the control group (placebo-control vs. 5-day treatment regimen) were used for dividing studies in different subgroups. Median and interquartile range (IQR) were converted to mean and SD according to the formulas suggested by Hozo et al (14). Besides, IQR of the 10-day arm in Goldman study was assumed equal to the 5-day arm due to the near median and first quartile between two arms.

## Results

From 329 records identified through database searching and references lists, 21 were selected for assessing full texts. After matching with our eligibility criteria, 5 studies were chosen for final inclusion (Figure1).The dosage of Remdesivir in these five studies was single 200mg intravenous dose in first day followed by once-daily 100mg intravenous dose from second day up to 10 days(15-19). Three multicenter randomized clinical trials were included in our study. Adaptive COVID-19 treatment trial (ACTT) and a trial in ten hospitals in china were both placebo-controlled and patients that were assigned to test arms received Remdesivir and those who were assigned to control arms received intravenous placebo with the same frequency and duration of Remdesivir infusions(17, 19). However, participants in both arms of third trial received Remdesivir but for different durations (18). 2 cohort studies were also included. Both of them assessed compassionate use of Remdesivir between 2 groups of patients: In Grein study, patients were divided based on the need for invasive oxygen support, while Antinori divided the patients into ICU and ward groups (15, 16). (Table 1 and 2) Inclusion criteria of the final studies are presented in table S1.

**Figure 1.**
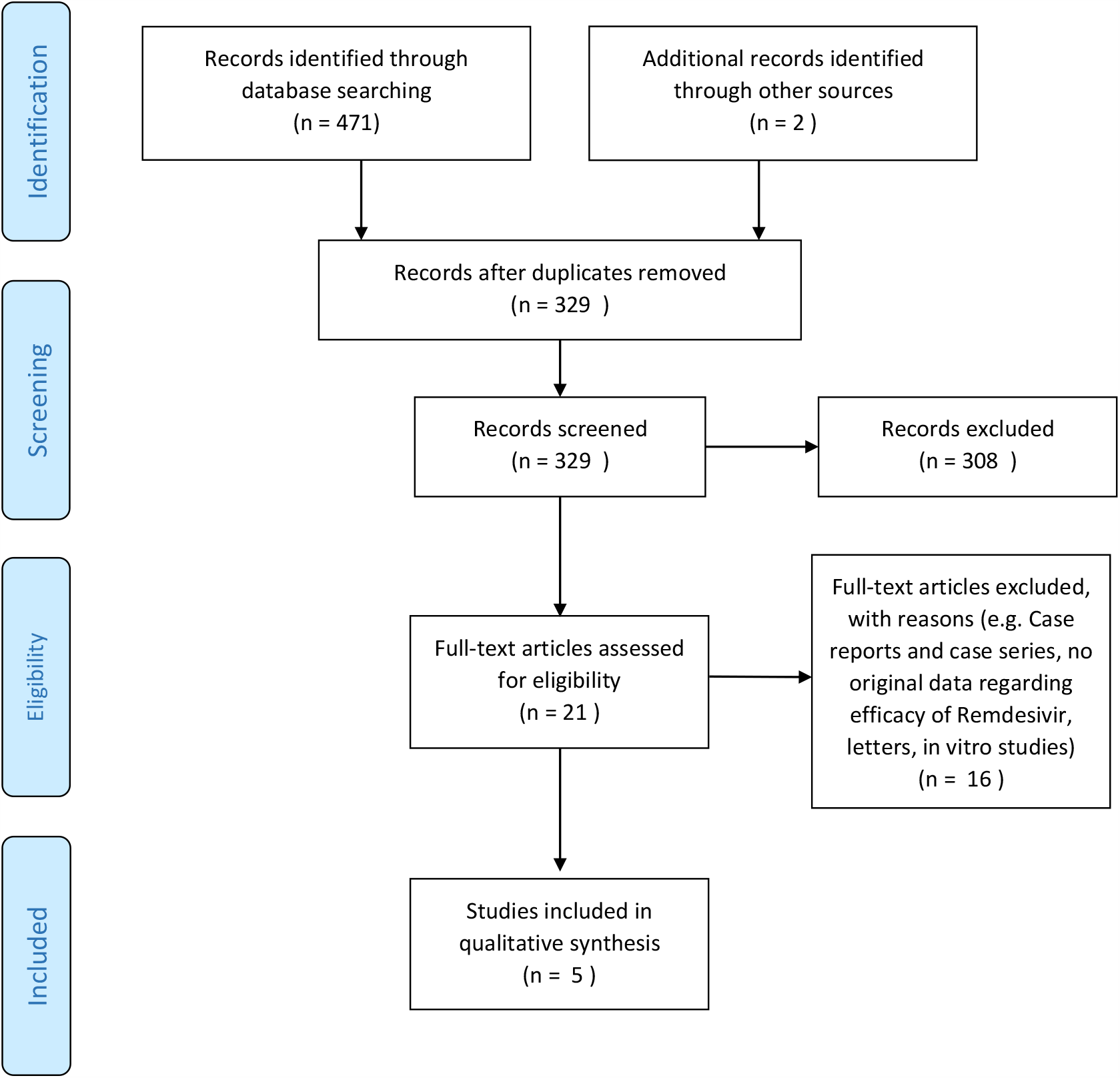
PRISMA flow chart.

**Table 1.** Details about study design and participants

| Authors | Type of study | Selected details of study design and participants | Group 1 sample size | Group 2 sample size | median duration of illness before RDV therapy |
| --- | --- | --- | --- | --- | --- |
| *Jonathan Grein, et al.* | Cohort study | Group1:patients needed Invasive oxygen support | 34 | 19 | 12days (IQR 9–15) |
|  |  | Group2:patients needed non-invasive oxygen support |  |  |  |
| *Spinello Antinori, et al.* | Cohort study | 31 patients had previously received LPV/r and HCQ | 18 | 17 | 7 days (IQR 5-10) * |
|  |  | Group1: 18 ICU patients |  |  | 4 days (IQR 3-5) ** |
|  |  | Group2: 17 ward patients |  |  |  |
| *Yeming Wang, et al.* | Double-blind, placebo-controlled RCT | Use of other medicines was permitted. | 158 | 79 | 10 day (IQR 9-12) |
|  |  | Group1 : test arm |  |  |  |
|  |  | Group 2 : control arm |  |  |  |
| *Beigel et al.* | Double-blinded, placebo-controlled RCT | Other specific treatment for COVID 19 were prohibited after enrollment, unless a written guideline implemented by hospital. | 538 | 521 | 9 days (IQR 6-12) |
|  |  | Group1 : test arm |  |  |  |
|  |  | Group 2 : control arm |  |  |  |
| *Jason D. Goldman, et al.* | Open-label, RCT | Group1 : 5-day arm | 200 | 197 | 8 days (IQR 5–11)† |
|  |  | Group2: 10 day arm |  |  | 9 days (IQR 6-12)†† |

Abbreviations: LPV/r - Lopinavir/Ritonavir, HCQ - Hydroxychloroquine, IQR - interquartile range, RDV – Remdesivir
\*median duration of illness before hospitalization
\*\*median duration from hospitalization to Remdesivir therapy
† median duration of illness before RDV therapy in 5-day arm
†† median duration of illness before RDV therapy in 5-day arm

**Table 2.**
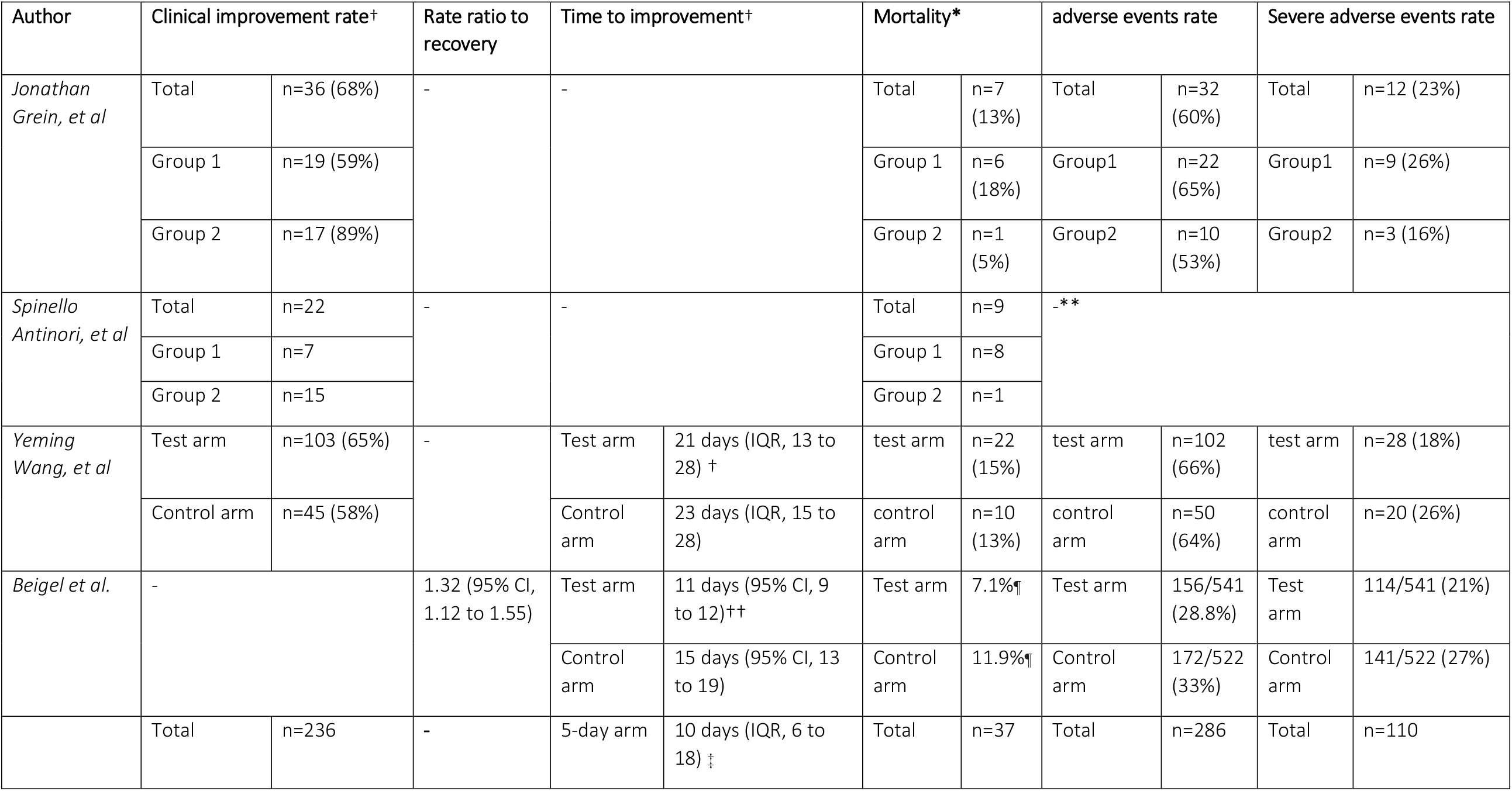
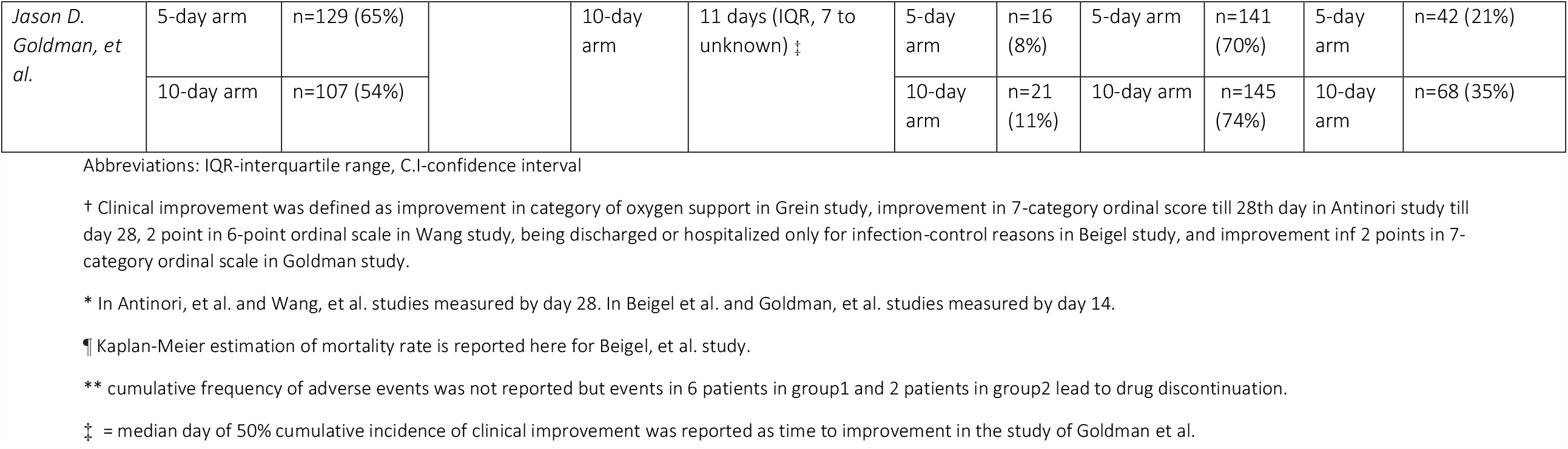
measured outcomes in the included studies

### RCT

In the preliminary report of ACTT, results of the analysis on 1059 participants with lower respiratory tract involvements of COVID-19 were published (17). In this double-blinded study, 77% of patients required oxygen administration at enrollment. The median time between illness onset and randomization of participants was 9 days (IQR 6 to 12) and no significant imbalances found in baseline characteristics of 538 participants in test arm and 521 patients in control arm. Primary outcome in this trial was time to recovery which defined as time from enrollment to the first day on which patient being discharged or hospitalization extended only for infection-control reasons. Time to recovery was 11 days in test arm (95% CI, 9 to 12) and 15 days in control arm (95% CI, 13 to 19). The rate ratio of recovery in 14th day was 1.32 (95% CI, 1.22 to 1.55) with no significant interaction with baseline clinical status. Mortality was higher in test arm; however, the difference was not statistically significant with or without adjustment of baseline illness severity. 21% of participants in test arm and 27% in control arm experienced serious adverse events. There are a number of considerations that must be taken into account when interpreting these results.

First, the patients were allowed to use other medications along with Remdesivir or placebo according to each hospital policy. This heterogeneity in medications across different subgroups of patients affects the generalizability of the results. Thus, the observed effects may be due to the combined effects of different medications and Remdesivir making the results less generalizable. Second, careful evaluation of the study tables revealed that only 180 out of 541 (33.3%) patients in the Remdesivir group and 185 out of 522 (35.4%) patients in the placebo group received the full 10 doses of complete treatment at the time of data analysis. Accordingly, the results may not be truly reflective of the response that would have been observed in the case of receiving the complete course of therapy.

Lastly, this trial is ongoing and preliminary results were published. Therefore, the analyses of safety and efficacy of Remdesivir in this study has not been completed yet and this imposes a high risk of bias. (Figure2)

**Figure 2.**
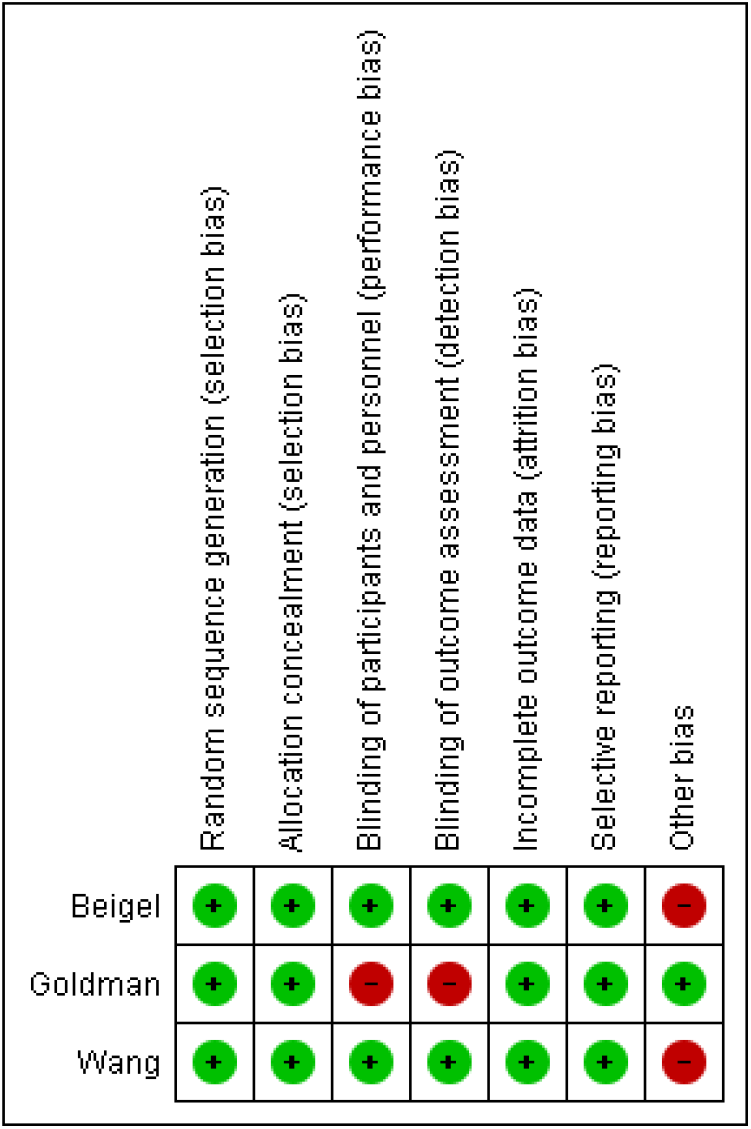
Risk of bias assessment of RCTs with Cochrane Collaboration’s Risk of Bias 2.0 tool for RCTs (ROB-2)

In a study on 237 patients with COVID-19 pneumonia and concomitant impaired oxygenation, the effect of Remdesivir on clinical improvement was not statistically significant (19). Median time to clinical improvement was 21 days (IQR 13 to 28) in test arm compared to 23 days (IQR 15 to 28) in control arm, and rate of clinical improvement by day 28 was 65% in test arm and 58% in control arm. Also, Remdesivir was not associated with a significant increase in negative conversion rate of viral load reduction in nasopharyngeal and sputum samples being compared to placebo. Researchers in this study calculated a sample size of 453 for 80% event rate within study duration of 28 days and 10% drop out rate, however, only 237 patients were enrolled.

There is a high risk of bias due to baseline characteristics imbalances resulted from inadequate sample size (Figure2); participants in test arm had more comorbidities, more tachypnea, and longer duration of illness. Participants of test arm had more adverse events (66% vs. 64%) and patients in control arm had more serious adverse events (26% vs. 18%).

An open-label RCT demonstrated that a 5-day course of Remdesivir, 200mg in first day and 100 mg for next 4 days, may be sufficient (18). In this trial, from 397 COVID-19 hospitalized patients, who did not require invasive ventilation at enrollment, 200 patients received 5-day course of Remdesivir and 197 patients received Remdesivir for 10 days. Randomization has taken place, but lack of stratification has led to worse baseline clinical status of 10-day arm patients. Rate of clinical improvement, which was defined as an improvement of more than one point on a 7-point ordinal scale by day 14, was 65% in 5-day arm patients and 54% in 10-day arm patients. Nevertheless, rate of clinical improvement, clinical recovery, discharge or mortality by day 14 were not significantly different between two arms after adjustment of baseline clinical status. However, it should be stated that among patients receiving invasive mechanical ventilation at day 5, ones assigned to 10-day (n=41) regime had lower mortality as compared to 5-day (n=25) regime (17% vs. 40%). In analysis of drug safety, both arms were statistically similar in incidence of adverse events; however, serious adverse events were more prevalent in 10-day arm patients after adjustment of baseline clinical status. Interpretation of Remdesivir efficacy based on this study is limited by the lack of placebo group.

### Cohorts

In a cohort study, Grein et al. analyzed the available data of 53 patients (15). Median duration of illness before Remdesivir therapy and median follow up time were 12 (IQR 9-15) and 18 days (IQR 13-23), respectively. Of 53 patients, 34 needed invasive oxygen support and the other 19 needed non-invasive oxygen support. 59% of patients in invasive oxygen support group had improvement in category of oxygen support as compared to 89% in patients in the other group by the end of the follow-up. Mortality rate was higher in invasive group (18% vs. 5%). Participants in invasive group experienced more adverse events (65% vs. 53%). For entering this study, patients’ physicians had to apply to receive Remdesivir. This limits the generalizability of the results. Besides, Lack of control group and insufficiency of follow up time are also other sources of bias in this study. (Table3).

**Table 3.** Risk of bias assessment of two cohort studies with Newcastle–Ottawa scale

| Author | Selection |  |  |  | Comparability | Outcome |  |  | Total |
| --- | --- | --- | --- | --- | --- | --- | --- | --- | --- |
|  | Representativeness | Selection of the non-exposed cohort | Ascertainment of exposure | Demonstration |  | Assessment of outcome | Follow-up | Adequacy of follow-up |  |
| Antinori | C | NA | A* | A* | C* | A* | A* | A* | 6 |
| Grein | C | NA | A* | A* | C* | A* | A* | C | 5 |

In the other cohort study of 35 patients, median duration of illness before hospitalization and median duration of admission before Remdesivir therapy were 7(IQR 5-10) and 4(IQR 3-5) days, respectively(16). 31 patients had been treated with Lopinavir/Ritonavir and Hydroxychloroquine previously, but after enrollment, Lopinavir/Ritonavir treatment was stopped. 9 of 18 ICU patients and 13 of 17 ward patients completed the 10-day course of Remdesivir therapy. ICU patients had lower clinical improvement rate (38.9% vs. 88.2%) by day 28. The median duration of viral load conversion in 22 patients with negative PCR tests was 12 days (IQR 9.25-16.75). The most common severe adverse events observed were elevation of liver enzymes (42.8%) and acute kidney injury (22.8%).

### Mortality

Visual inspection of figure 3 showed that mortality rates were higher in cohort studies compared to RCTs. It suggested the lower mortality rates in controlled situations of RCTs compared to real-world situation in cohort studies. Besides, Remdesivir efficacy in reducing 28-day mortality rate was not significantly lower than its efficacy in reducing 14-day mortality rate (figure 4). Forest plot of mortality rates in placebo-control and 10-day vs. 5-day studies is presented in figure S1.

**Figure 3.**
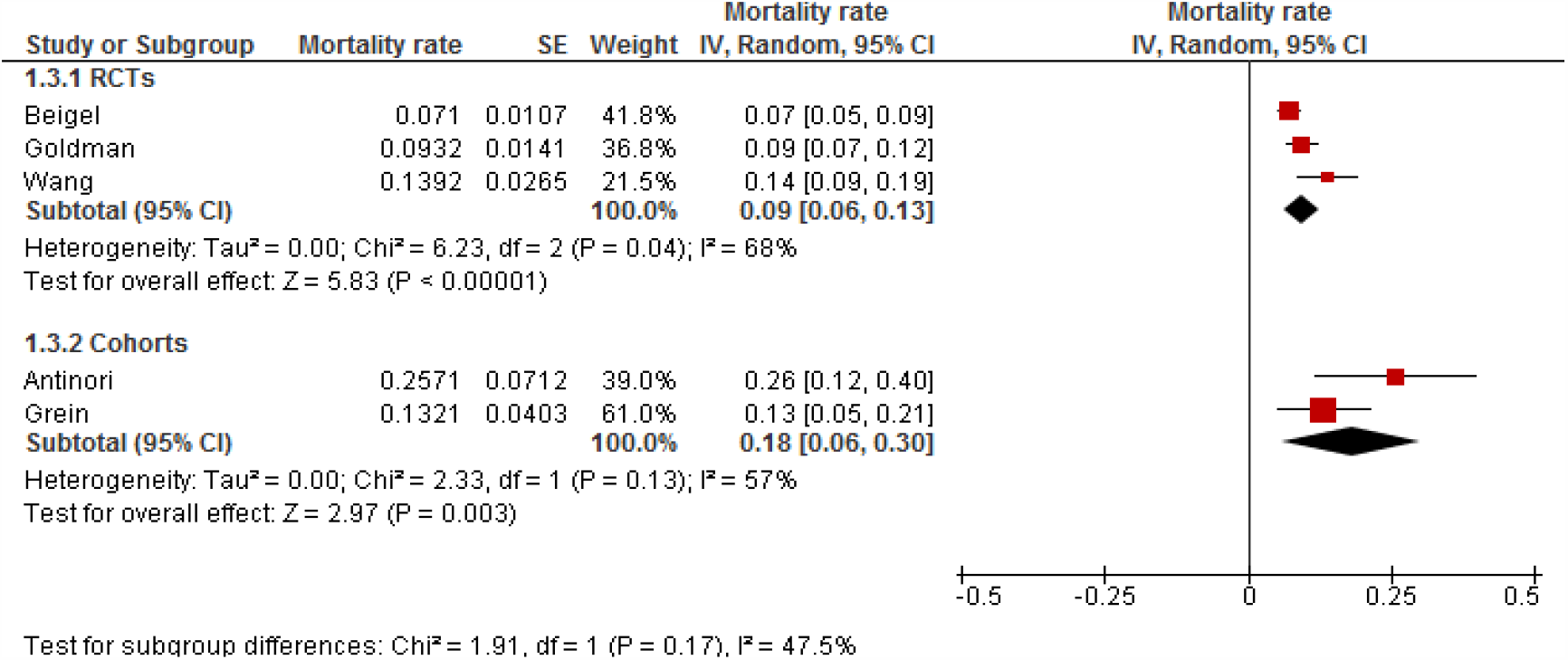
Forest plot of mortality rates in placebo-control and 10-day vs. 5-day studies.

**Figure 4.**
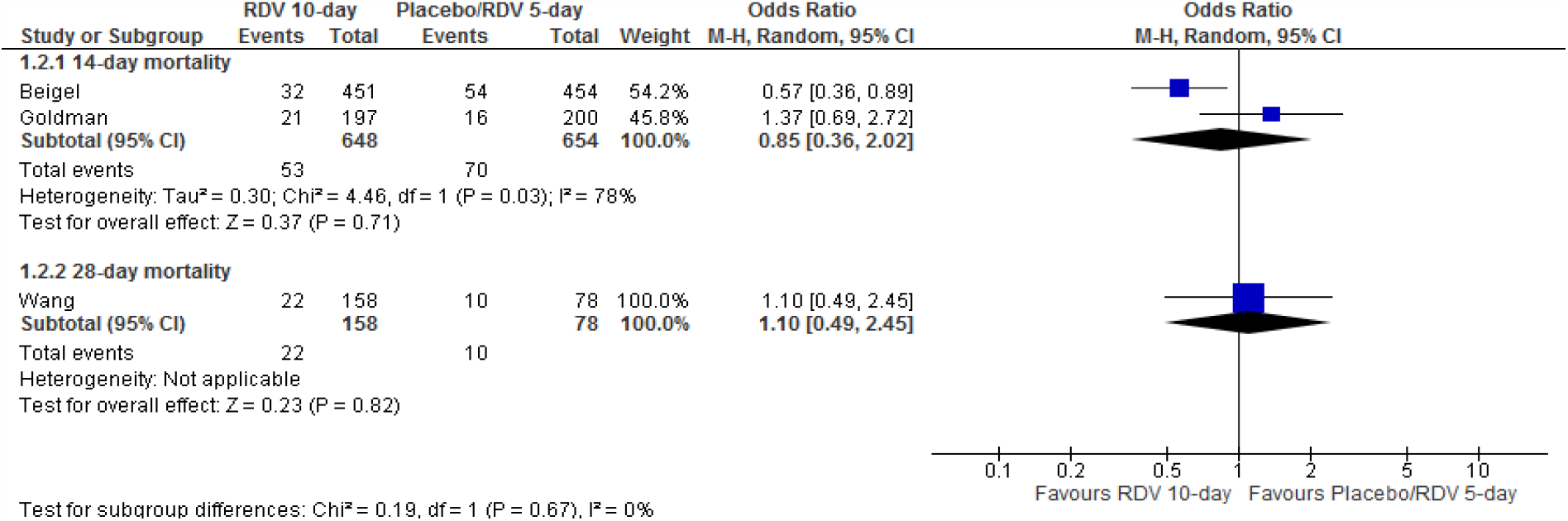
Forest plot of 14-day vs. 28-day mortality rates in RCTs.

### Clinical improvement

As demonstrated in figure 5, improvement rates in RCTs and cohorts were not significantly different, contrary to mortality rates. As was seen in mortality rate, Remdesivir efficacy in increasing improvement rate was not significantly different by day 14 and day 28 (figure 6). Figure S2 shows improvement rates in placebo-control and 10-day vs. 5-day studies.

**Figure 5.**
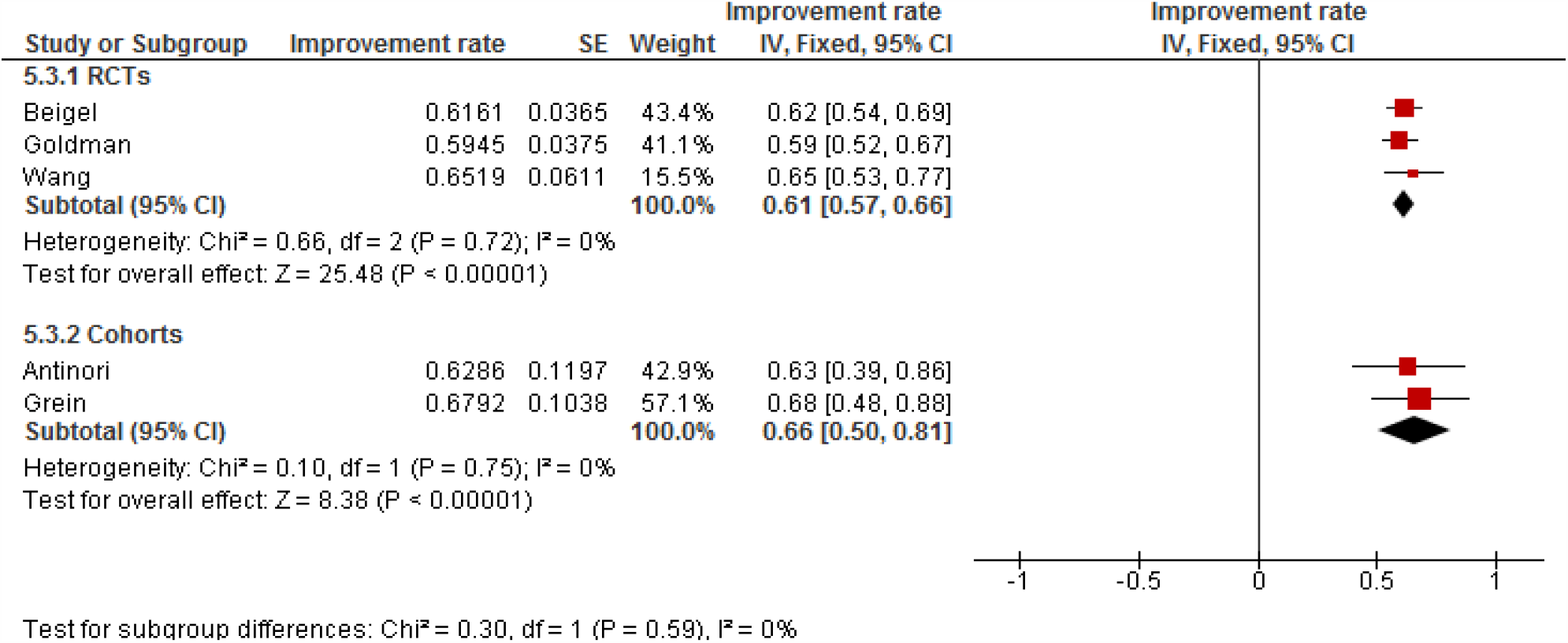
Forest plot of mortality rates in placebo-control and 10-day vs. 5-day studies.

**Figure 6.**
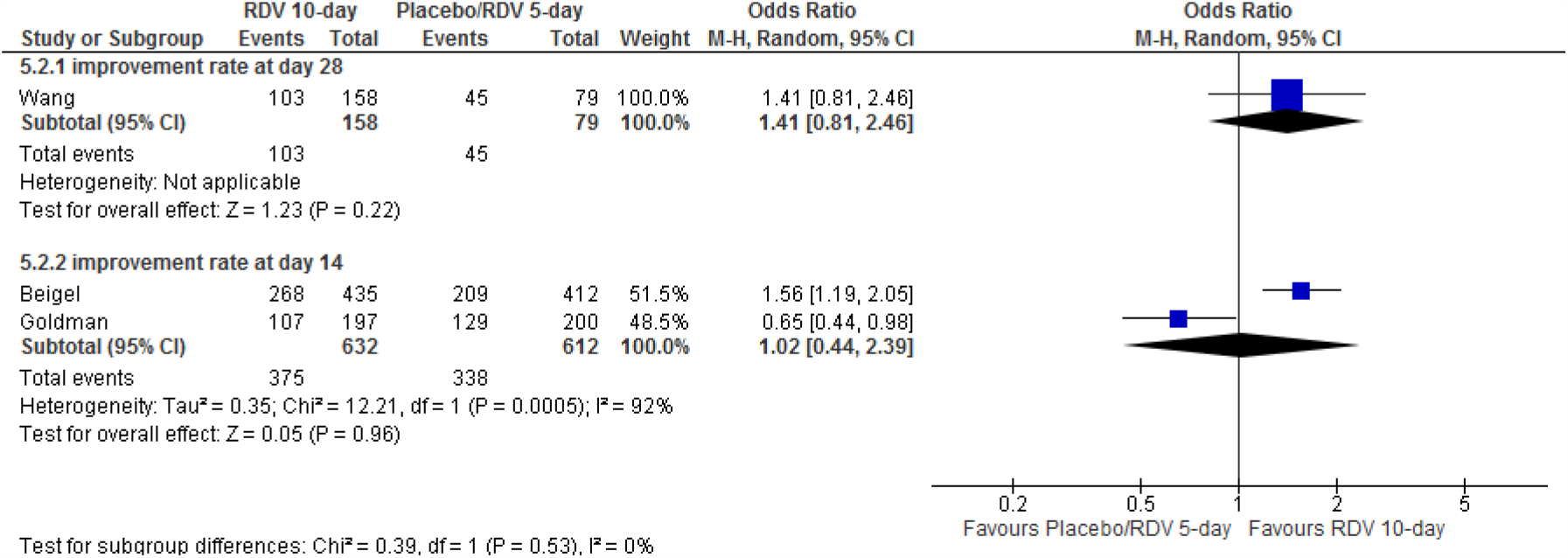
Forest plot of 14-day vs. 28-day improvement rates in RCTs.

### Time to clinical improvement

Figure 7 showed Remdesivir effects on reducing the time to clinical improvement. It showed that 5-day regimen can reduce time to clinical improvement compared to placebo, and continuing Remdesivir for another 5 days can even cause more decrease in time to improvement than 5-day regimen.

**Figure 7.**
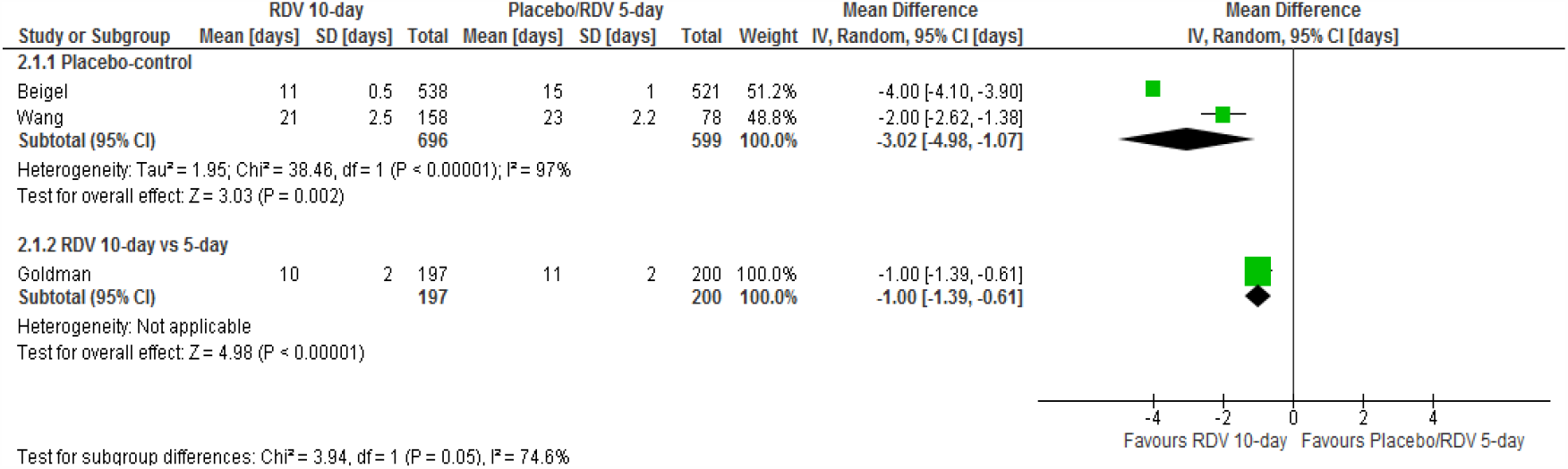
Forest plot of time to clinical improvement rates in placebo-control and 10-day vs. 5-day RCTs.

### All adverse events

Patients received 10-day Remdesivir therapy showed lower adverse events compared to placebo-control group. Besides, patients with 5-day Remdesivir regimen experienced lower adverse event rate than the 10-day group. However, both of the above comparisons did not show significant differences between two groups. Furthermore, superiority of the 5-day arm to the placebo arm is not notable due to the remarkable overlap between 95% C.I of the odds ratio (figure 8). Forest plot of all adverse event rates in RCTs and cohort studies is presented in figure S3.

**Figure 8.**
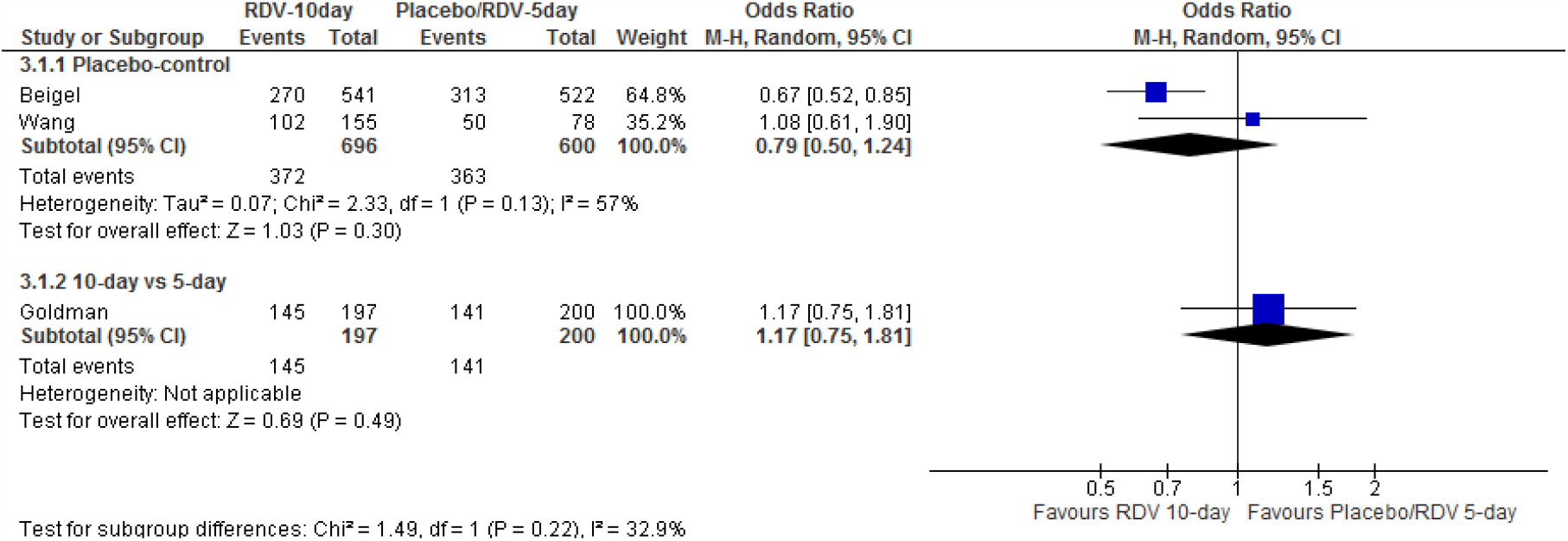
Forest plot of all adverse event rates in placebo-control and 10-day vs. 5-day RCTs.

### Severe adverse events

10-day regimen showed lower severe adverse events rate than the placebo-control group. In addition, 5-day regimen group had lower adverse event rate compared to the 10-day group. Contrary to all adverse events, superiority of the 5-day regimen to the 10-day regimen and 10-day regimen to the placebo group are significant for severe adverse events (figure 9). Figure S4 shows severe adverse event rates in RCTs and cohort studies.

**Figure 9.**
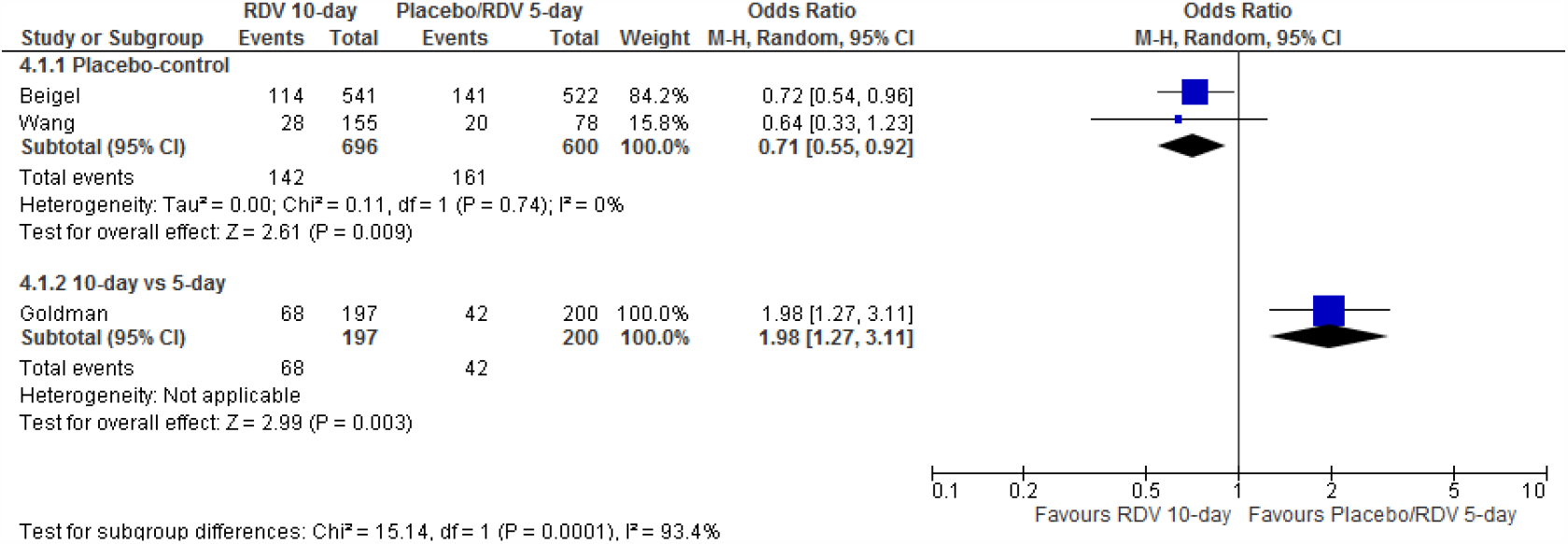
Forest plot of severe adverse event rates in placebo-control and 10-day vs. 5-day RCTs.

## Discussion

After searching for all the manuscripts evaluating the use of Remdesivir in the clinical management of COVID-19, 3 RCTs and 2 cohort studies were identified. Unfortunately, the heterogeneity in study design and definition of clinical outcomes as well as controversial study results make it hard to establish a solid conclusion regarding the clinical efficacy of Remdesivir. ACTT reported that Remdesivir is useful in reducing the time to recovery in patients with COVID-19 pneumonia (17). On the other side, Wang et al. failed to show a significant benefit associated with the use of Remdesivir in terms of clinical outcomes (19). Goldman study revealed that 5-day and 10-day Remdesivir therapy does not have significant differences in the terms of clinical outcomes.(18) Finally, it is suggested in 2 cohort studies that the use of Remdesivir may improve clinical outcomes; although, their results must be interpreted carefully considering the lack of a control group and incomplete understanding of the natural course of the virus (15, 16).

To date, Beigel et al. have published the largest RCT evaluating the use of Remdesivir in patients with lower respiratory tract involvement (17). Large sample size and a well-designed study protocol are the strengths of this study. Remdesivir group was superior to the placebo group reducing the time to recovery and increasing rate of recovery. Based on this study, Remdesivir is safe, however, patients with severe renal impairments were excluded.

The report of a RCT in 10 hospitals in china demonstrates no benefit in clinical outcomes in using Remdesivir for treatment of COVID-19 patients with lower respiratory tract involvement (19).

However, the inability to recruit the predetermined study population resulted in study power reduction from 80% to 58%, as Wang et al. stated in the manuscript. Furthermore, the imbalances in baseline characteristics of Remdesivir and placebo groups were also a source of error in this study. Low study power and higher severity of illness in Remdesivir arm both decreases the probability of detecting Remdesivir effectiveness. For example, a separate analysis in a subgroup of patients with less than 10 days from symptom onset to the start of treatment showed lesser time to clinical improvement 1.52 (95% CI, 0.95 to 2.43)and faster rate of sputum viral load reduction in the Remdesivir group (0.0672). Mentioned outcomes could be statistically significant if this study had higher power and was able to detect smaller effect. As a whole, this study was not in favor of using Remdesivir to treat COVID-19 patients; although, there was an underestimation toward Remdesivir effectiveness.

Goldman et al. study showed that 5-day course and 10-day course of Remdesivir therapy did not have significant difference (18). Patients receiving mechanical ventilation or extracorporeal membrane oxygenation were not enrolled in this study. Therefore, less severe patients were evaluated in this study. Unlike the 2 other RCTs, assessing the efficacy of Remdesivir is impossible due to the lack of the placebo control group. Baseline clinical characteristics of the patients in two arms were different because of the absence of stratification. After adjustment of baseline imbalances, both 5-day and 10-day groups showed near outcomes. Although, severe adverse events were more prevalent in the 10-day group. Furthermore, in patients requiring invasive mechanical ventilation after receiving Remdesivir for 5 days, continuation of therapy for another 5 days was associated with lower mortality rate. Due to the shortage of Remdesivir supply (20), indifference between outcomes of these 2 regimes helps us to treat more severe patients.

Grein et al. stated that Remdesivir may be useful in treating severe COVID-19 patients (15). 64% of the patients needed invasive mechanical ventilation and the sample was correctly labeled as severe. This study showed that clinical improvement in milder cases were higher than that of more severe cases. It prompts the need for earlier recognition and initiation of treatment before the disease advances to severe stages. However, the results may be confounded by several reasons. First, receiving previous treatment and differences in length of Remdesivir therapy may affect the final results. Second, absence of a control group to compare the results with, make it difficult to understand whether the reported improvement rate is due to Remdesivir or not. Third, authors in this study considered death as a censoring event. Reanalysis of the data for correcting this error, showed that estimated improvement rate by Kaplan-Meier analysis falls from 84% to 70%. Lastly, by considering 32 patients who are discharged or died by the end of the study as the denominator, mortality rate rises from 13% to 22% (21).

A study of Remdesivir therapy in 35 ICU and ward patients showed that ward patients benefit more from Remdesivir therapy than ICU patients and experience fewer adverse events. Also, this study reminds the higher efficacy of Remdesivir therapy in less severe patients.

When we considered mortality as the outcome of interest, effectiveness of Remdesivir in treating COVID-19 patients was higher than its efficacy; however, this superiority was not observed when improvement rate was chosen as the target outcome. Besides, as mentioned previously, Remdesivir efficacy in reducing mortality rate and increasing improvement rate by day 28 and day 14 are not much different. Therefore, we suggested that most benefits from Remdesivir therapy appear in the first 14 days from the start of the treatment and final conditions of the patients are mostly determined by day 14. Also, we found that extending Remdesivir treatment to 10 days lead to the faster clinical improvement.

All adverse events rates were not significantly different between 10-day arm, 5-day arm, and placebo groups. However, severe adverse events rates were lower in 5-day group compared to 10-day group, and the 10-day group showed lower severe adverse events compared to placebo group.

It is difficult to answer the question regarding the efficacy of using Remdesivir in severe COVID-19 patients. So far, there are only 2 studies comparing the efficacy of Remdesivir in test arm and placebo control arm, and the results of these two studies were controversial. Beigel et al. study results were in favor of using Remdesivir. While, Wang et al. study could not find significant benefit from Remdesivir therapy. We think that Remdesivir can be considered as a choice, especially in patients without invasive mechanical ventilation (17) or in patients with less duration of illness (19). The previous studies revealed that Remdesivir is beneficial; however, the magnitude of this benefit is not large enough to make Remdesivir monotherapy as an ultimate treatment. Further RCT studies with similar study designs and large sample sizes evaluating the efficacy of Remdesivir as a monotherapy and in combination with other choices should be conducted. Moreover, assessing Remdesivir efficacy in patients with different clinical subgroups and duration of symptoms are required for optimizing patient selection.

Our study has three major limitations. Firstly, only five articles were eligible to enter our study, at least three of them (15, 16, 19) did not have enough sample sizes in different clinical subgroups. Therefore, comparing these subgroups regarding the efficacy of Remdesivir was not incontrovertible. Second, pooling the data of the original articles was impossible due to heterogeneity in study design and reported outcomes. However, critical review of the study settings and methods and their associations with the reported results have enable us to compare the differences observed in results of these studies. Third, lack of control arm in 2 cohort studies(15, 16) and differences in baseline characteristics between test and control arms in 2 RCTs(17, 19), made it difficult to suggest a conclusive statement for the efficacy of Remdesivir in treating COVID-19.

## Conclusion

Based on the current evidence, 5-day course of Remdesivir therapy in COVID-19 patients is probably efficacious and safe. Remdesivir efficacy differs in different disease severity subgroups and hospitalized patients without invasive mechanical ventilation benefit the most from Remdesivir. Treatment can be extended to 10 days if satisfactory improvement is not achieved by day 5. Most benefits from Remdesivir therapy take place in the first 14 days, and patients conditions usually do not change dramatically in the next 14 days. More studies are needed to explore the efficacy of Remdesivir monotherapy or combination therapy in different disease severity subgroups.

## Data Availability

The original data will be available on request.

